# The signature features of COVID-19 pandemic in a hybrid mathematical model - implications for optimal work-school lockdown policy

**DOI:** 10.1101/2020.11.02.20224584

**Authors:** Teddy Lazebnik, Svetlana Bunimovich-Mendrazitsky

## Abstract

**Background:** The coronavirus disease 2019 (COVID-19) first identified in China, spreads rapidly across the globe and is considered the fastest moving pandemic in history. The new disease has challenged policymakers and scientists on key issues such as the magnitude of the first-time problem, the susceptibility of the population, the severity of the disease, and its symptoms. Most countries have adopted “lockdown” policies to reduce the spatial spread of COVID-19, but they have damaged the economic and moral fabric of society. Timely action to prevent the spread of the virus is critical, and mathematical modeling in non-pharmaceutical intervention (NPI) policy management has proven to be a major weapon in this fight due to the lack of an effective COVID-19 vaccine.

**Methods:** We present a new hybrid model for COVID-19 dynamics using both an age-structured mathematical model and spatio-temporal model in silico, analyzing the data of COVID-19 in Israel. The age-structured mathematical model is based on SIRD two age-class model. The spatial model examines a circle of day and night (with one-hour resolution) and three main locations (work / school or home) for every individual.

**Results:** We determine mathematically the basic reproduction number *R*_0_ via the next-generation matrix based on Markov chain theory. Then, we analyze the stability of the equilibria and the effects of the significant differences in infection rates between children and adults. Using the *hybrid* model, we have introduced a method for estimating the reproduction number of an epidemic in real time from the data of daily notification of cases. The results of the proposed model are confirmed by the Israeli Lockdown experience with a mean square error of 0.205 over two weeks. The model was able to predict changes in *R*_0_ by opening schools on September 1, 2020, resulting in *R*_0_ = 2.2, which entailed a month’s quarantine of all areas of life. According to the model, by extending the school day to 9 hours, and assuming that children and adults go to school and work every day (except weekends), we get a significant reduction in *R*_0_ of 1.45. Finally, model-based analytical-numerical results are obtained and displayed in graphical profiles.

**Conclusions:** The use of mathematical models promises to reduce the uncertainty in the choice of “Lockdown” policies. Our unique use of contact details from 2 classes (children and adults), the interaction of populations depending on the time of day (the cycle of day and night), and several physical locations, allowed a new look at the differential dynamics of the spread and control of infection. Using knowledge about how the length of the work and school day affects the dynamics of the spread of the disease can be useful for improving control programs, mitigation, and policy.

**Author summary:** Everybody in the modern world understands today that the pandemics threat is not less dangerous than the wars. COVID-19 showed us that pandemics effects the economies of all the countries over the world brings to a total lockdown of social life and enormous mortality. There was no effective vaccine/treatment, to stop the spread of COVID-19 and, therefore, policymakers have taken unprecedented measures, including quarantines, public health measures, travel bans, and others, without knowing in advance the effect of these restrictions.

In this study, we develop a mathematical model of the pandemic spread taking into account the different dynamics of the disease in two age groups of children and adults. Using this model we succeeded to simulate the COVID-19 spread in Israel. The current study accurately predicts the effect of the work/school lockdown on the outbreaks. We have proven that by keeping schools open and increasing the school day to 8-9 hours, infection rates are reduced. Our results also show that if at least half of the adult population is a lockdown, the effect of children’s isolation on the infection rate is small, indicating the importance of multiple age groups of the population in the selection of restrictions.

## Introduction and related work

At the beginning of 2020, the novel severe acute respiratory syndrome coronavirus 2 (SARS-CoV-2), also known as COVID-19 reached Europe and the western world from China [1]. Similar to other diseases from the coronavirus family, COVID-19 is transmitted human-to-human, but it turned out that COVID-19 is more infectious and transmissible than previous coronavirus [2].

The World Health Organization (WHO) has declared COVID-2019 a public health emergency of international concern [3,4]. Currently, due to a lack of an efficient vaccine or clinical treatment to COVID-19, policy-makers are forced to rely on non-pharmaceutical intervention (NPI) policies to reduce the infection rate and control the epidemic. A few examples of NPI policies are masks, social distancing, work capsules, and partial to a full lockdown of central locations (restaurants, malls, offices, etc.). These policies reduce the infection rate but have an influence on the economy, healthcare system, and social life.

Multiple studies have been conducted to study epidemics in general and the COVID-19 epidemic in particular from both biological and epidemiological perspective providing the information about the epidemics behavior with relevant data for later modeling [1, 5–9]. In addition, these studies examine the clinical characteristics of COVID-19, enabling a deeper understanding of the disease spreads in the population.

Mathematical models are shown to be a useful tool for policy-makers to make data-driven decisions based on investigation of different scenarios and their outcomes in a controlled manner [5, 10, 11]. These mathematical models can be divided into two main groups.

First, models aim to predict the different parameters such as the total COVID-19 related death and peak in hospitalized individuals, given the historical data up to some point. For example, Nesteruk [12] used the data from January 16 to February 9 (2020), from mainland China with the continuous SIR (S-susceptible, I-infected, R-recovered) model. Nesteruk [12] fitted the SIR model using the least mean square method, which resulted in poor fitting to later officially confirmed infected cases [4]. This can be explained by the fact that the SIR model with no modification is too simplistic to correctly represent the dynamics of the COVID-19 epidemic. In [12], a simple calculation method was proposed that allows quick results which can predict the COVID-19 spread with a fine accuracy if calculated using a very accurate data of infections and recoveries.

Model proposed by Tuite et al. [11] which used data from January 25 to March 1 (2020) Ontario, Canada, is SEIR (E-exposed) model, the extension of the SIR model. Authors of [11] took into consideration four levels of infection severity, social isolation in the exposed and infected states, hospitalization dynamics, and death state. [11] estimated that 56% of the Ontario population would be infected over the course of the epidemic with a peak of 55500 cases in intensive care units (ICU) and 107000 total cases. At the same time, in all of Canada there are only half of this number of infections [4]. This error in their prediction is associated with incomplete information due to lower frequency of tests and inaccurate ICU records. Nevertheless, Tuite et al’s [11] model presents a more detailed dynamic between the infected population and the healthcare response which policy-makers can take into consideration.

Ivorra et al. [13] proposed nine states model divides individuals into isolated, hospitalized, or dead. In addition, authors of [13] take into consideration the fact that the recorded confirmed data is under-sampled.

Furthermore, a machine learning-based model was proposed by Allam et al. [14] using several machine learning algorithms (Knn, Random Forest, Support Vector Machine) on data of 53 clinical cases. This model was used to predict infection severity and spread. However, this approach suffers from an imbalanced sample of the distribution of severity in the population, as simple or asymptomatic cases are not reported, leading to poor representation of the real dynamic.

Second, models aim to analyze and optimize NPI policies. A model proposed by Zhao et al. [15] is an extension to the SEIR model where the susceptible population is separated into two groups: individuals not taking infection-prevention actions and people taking infection-prevention actions as an NPI policy. In addition, authors of [15] included a probability of willingness to take infection-prevention actions with changes over time. They predicted 148.5 thousand infections by the end of May 2020 in Wuhan alone while in all of China there were 84.5 thousand infections at the same time [4]. The authors introduces a stochastic element to the SEIR model making it more robust for social changes happened during the epidemic.

Di Domenico et al. [16] used data from March 17 to May 11 (2020) Île-de-France with a stochastic age-structured transmission extension of the SEIR model integrating data on age profile and social contacts of four age-based classes. In this model, hospitalization dynamics with ICU cases are taken into consideration in the model. Model shows that during full lockdown the reproductive number is estimated to be 0.68, due to an 81% reduction of the average number of contacts. These results show that dividing the population into several age-based classes better represent the spread of COVID-19 from an epidemiological perspective [17, 18].

In this paper, we provide and study a more accurate spatio-temporal model for the COVID-19 transmission by using individual two age classes SIRD named *hybrid* model (D-death) model. We study two important factors concerning the diffusion of COVID-19: schooling/working hours and physical location of infected population has provided new insights into the epidemic dynamics. Based on the different impact of COVID-19 to the immune response, severity of infection and transmission disease in different age groups (mainly children and adults) [17, 19], we proposed a two classes age-structured SIRD epidemic model dividing the population into children and adults. Moreover, we developed a numerical, stochastic simulator based on this *hybrid* model (https://teddylazebnik.info/coronavirus-sir-simulation/index.html) for COVID-19 population spread in addition to the analytical examination of the epidemic dynamics.

This paper is organized as follows: First, we introduce our mathematical *hybrid* model based on the SIRD model with dual age-structured. Second, we present the model’s equilibria, stability analysis, and asymptotic form. Third, we present the spatial model extending the SIRD model by introducing a day-night circle and three locations of disease transmission (home, work, school). Fourth, an analysis of NPI policies including optimal lockdown, optimal work-school duration is presented, and a comparison to the Israeli historical data from August and September. Finally, we discuss the main epidemiological results arising from the model.

## Hybrid model

As shown in [20] and [21], the spatial model plays an important role in describing the spreading of communicable diseases, because individuals move around inside a zone or habitat in time.

In this section, we present a *hybrid* model (Fig 1) that is based on a SIRD model for two age classes using eight populations (dynamics are shown in Fig 2) and a spatial model where these populations are distributed in space and time between work, school and home (Fig 5). We will define these sub-models in the following sections.

**Fig 1.**
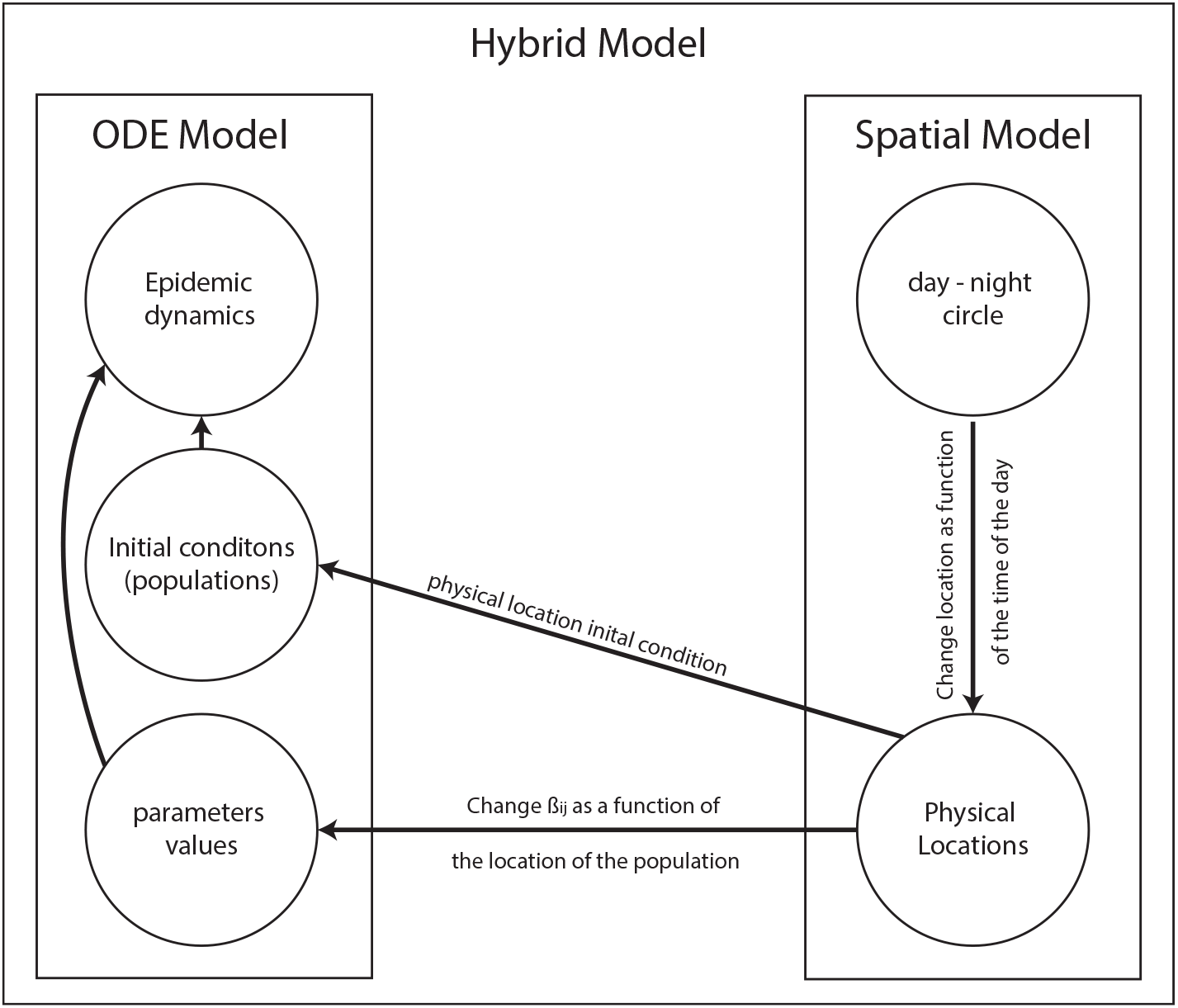
Schematical view of the *hybrid* model’s components and relationship between the ODE and the spatial models.

**Fig 2.**
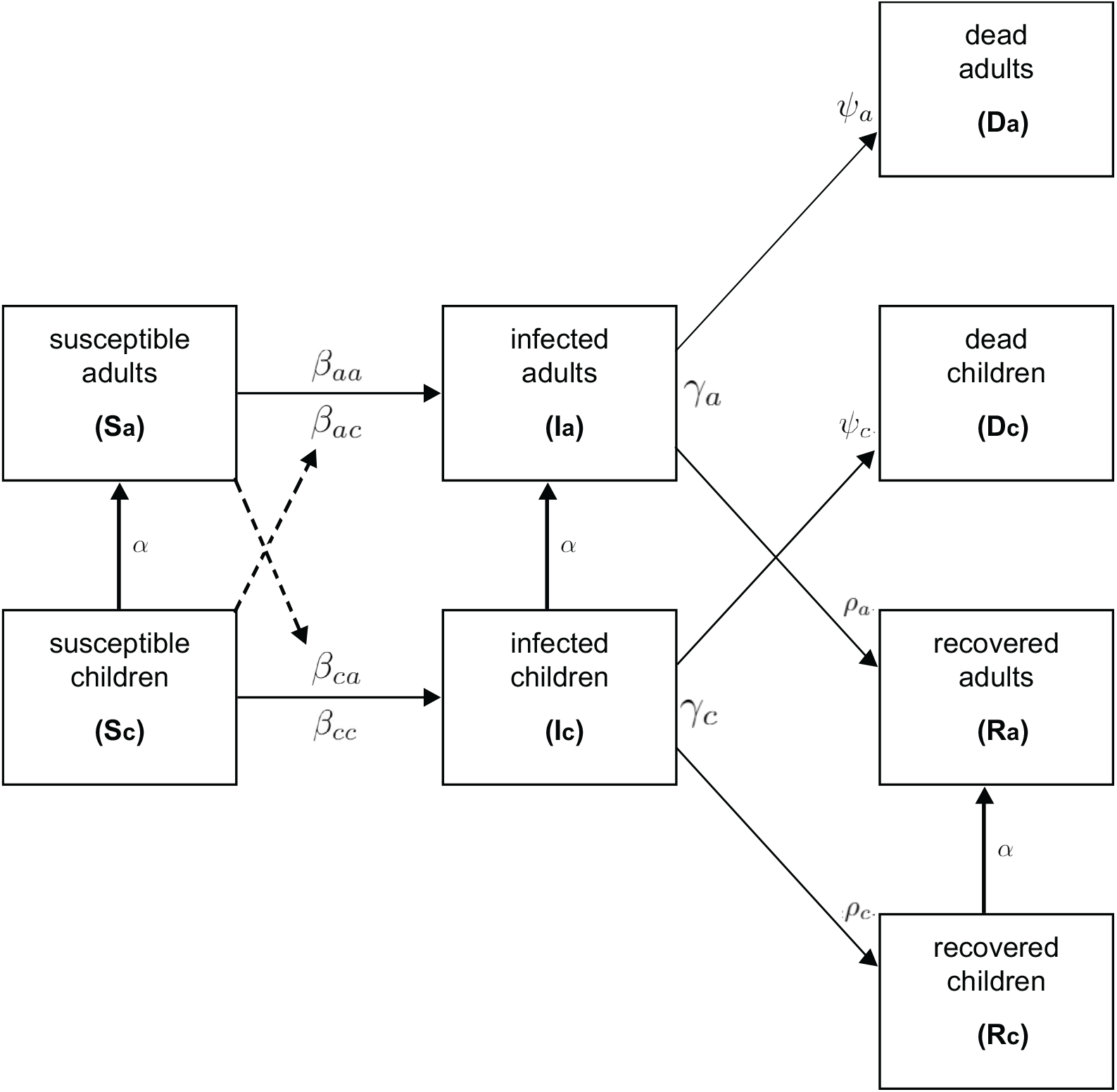
Schematical view of transition between disease stages, divided by age-class.

### Two class age-structured epidemic model

The SIR model is proven to be a meaningful mathematical tool for epidemic analysis [22]. This model with the needed modifications has already been shown to predict the epidemics such as COVID-19 [23], influenza [24], Ebola [10], and others.

Data from several epidemiological studies show that children and adults transmit the disease at different rates [17, 19]. In addition, adults, on average, have a much longer recovery duration compared to children [6, 8]. Therefore, a SIR model which takes into consideration the different age groups better represents the epidemiological population dynamics. An extension of the SIR model to two age-classes has been investigated for explanation of Polio outbreak by Bunimovich-Mendrazitsky and Stone [25].

### Model definition

The model considers a constant population with a fixed number of individuals *N*. Each individual belongs to one of the three groups: susceptible (*S*), infected (*I*), and recovered (*R*) such that *N* = |*S*| + |*I*| + |*R*|. When an individual in the susceptible group (*S*) exposed to the infection, it is transferred to the infected group (*I*). The individual stays in this group on average *d*_*I→R*_ days, after which it is transferred to the recovered group (*R*).

In addition to these groups, we define a death group (*D*) which is associated with individuals that are not able to fully recover from the disease or succumb to death. Individuals from the infected group (*I*) recover from the disease in some chance and move to (*R*), while the others do not and move to (*D*). Therefore, in each time unit, some rate of infected individuals recover while others die or remain seriously ill.

We divide the population into two classes based on their age: children and adults because these groups experience the disease in varying degrees of severity and have different infection rates. In addition, adults and children are present in various discrete locations throughout many hours of the day which affects the spread dynamics. Individuals below age *A* are associated with the *“children”* age-class while individuals in the complementary group are associated with the *“adult”* age-class. The specific threshold age (*A*) may differ in different locations but the main goal is to divide the population into two representative age-classes. Since it takes *A* years from birth to move from a child to an adult age group, the conversion rate is set as 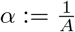.

By expanding the designation to two age-classes, we let *S*_*c*_, *I*_*c*_, *R*_*c*_, *D*_*c*_ and *S*_*a*_, *I*_*a*_, *R*_*a*_, *D*_*a*_ represent susceptible, infected, recovered, and death groups for children and adults, respectively such that

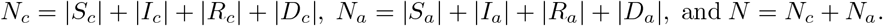

The model does not take into consideration death during the epidemic unrelated to the disease itself because in the United States in 2018 the birth rate was 11.6 for every 1000 individuals [26] while the mortality rate in 2017 was 8.6 for every 1000 individuals [27], resulting in around 0.3% increment of the population size which is assumed to be small enough to be neglected. In addition, we introduce two death states for children and adults, respectively, that died from the epidemic.

Eq (1-10) describes the epidemic’s dynamics.

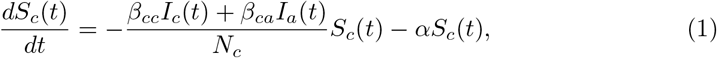

In Eq (1), 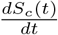 is the dynamical amount of susceptible individual children over time. It is affected by the following three terms. First, with rate *β*_*cc*_ each infected child infects susceptible children. Second, with rate *β*_*ca*_ each infected adult infects the susceptible children. Finally, children grow and pass from the children’s age-class to the adult’s age-class with transition rate *α*, reduced from the children’s age-class. *N*_*c*_ is the size of the children population and used to take all variables as fixed proportions of the population *N*.

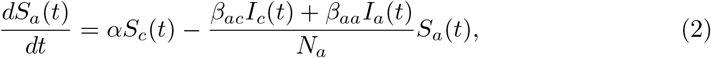

In Eq (2), 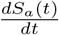 is the dynamical amount of susceptible adult individuals over time. It is affected by the following three terms. First, children grow and pass from the children’s age-class to the adult’s age-class with transition rate *α*, added to the adult age-class. Second, with rate *β*_*ac*_ each infected child infects the susceptible adult. Finally, with rate *β*_*aa*_ each infected adult infects a susceptible adult. *N*_*a*_ is the size of the adult population and used to take all variables as fixed proportions of the population *N*.

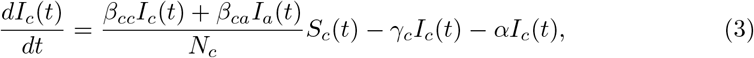

In Eq (3), 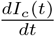 is the dynamical amount of infected individual children over time. It is affected by the following four terms. First, with rate *β*_*ca*_ each infected child infects the susceptible adult. Second, with rate *β*_*cc*_ each infected child infects a susceptible child. Third, individuals recover or die from the disease after a period *γ*_*c*_. Finally, children grow and pass from the children’s age-class to the adult’s age-class with transition rate *α*, reduced from the adult’s age-class. While the last process has a minor impact relative to the first three processes, we do not neglect it in order to count edge-cases.

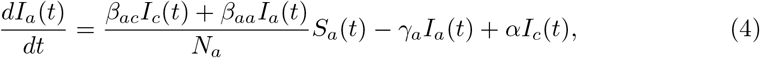

In Eq (4), 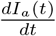 is the dynamical amount of infected adult individuals over time. It is affected by the following four terms. First, with rate *β*_*ac*_ each infected child infects the susceptible adult. Second, with rate *β*_*aa*_ each infected adult infects a susceptible adult. Third, individuals recover or die from the disease after period *γ*_*a*_. Finally, children grow and pass from the children’s age-class to the adult’s age-class with transition rate *α*, added to the adult’s age-class. While the last process has a minor impact relative to the first three processes, we do not neglect it to count edge-cases.

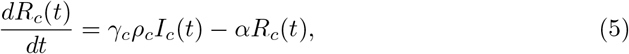

In Eq (5), 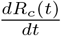 is the dynamical amount of recovered individual children over time. It is affected by the following two terms. First, in each point, a portion of the infected children recover after period *γ*_*c*_ which is multiplied by the rate of children that do recover from the disease *ρ*_*c*_. Second, children grow from birth and pass from the children’s age-class to the adult age-class with transition rate *α*, reduced from the children’s age-class.

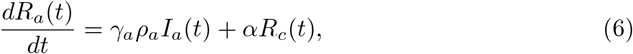

In Eq (6), 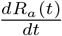 is the dynamical amount of recovered adult individuals over time. It is affected by the following two terms. First, in each point, a portion of the infected adults recover after period *γ*_*a*_ which is multiplied by the rate of adults that do recover from the disease *ρ*_*a*_. Second, children grow from birth and pass from the children’s age-class to the adult age-class with transition rate *α*, added to the adult age-class.

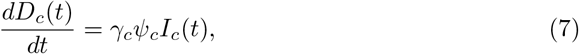

In Eq (7), 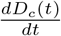 is the dynamical amount of dead individual children over time. It is affected by the portion of the infected children that do not recover after period *γ*_*c*_ which is multiplied by the rate of children that do not recover from the disease *ψ*_*c*_.

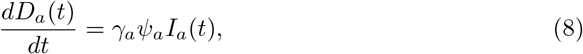

In Eq (8), 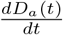 is the dynamical amount of dead adult individuals over time. It is affected by a portion of the infected adult that do not recover after period *γ*_*a*_ which is multiplied by the rate of adults that do not recover from the disease *ψ*_*a*_.

It should be noted that both Eq (7) and (8) can be presented as one equation

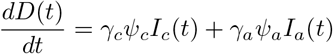

as *D*_*a*_ and *D*_*c*_ are accumulative populations which do not infect the dynamics separately. Nevertheless, to provide a better representation of the age-based death in the epidemic, we chose to divide the dead individuals into the same age-groups which allows later analysis of the death of children and adults separately. In summary, the interactions between disease stages presented in Fig 1 are modelled by the following system of eight coupled ordinary differential equations (ODE) Eq (9) with initial conditions at *t* = 0, Eq (10) as:

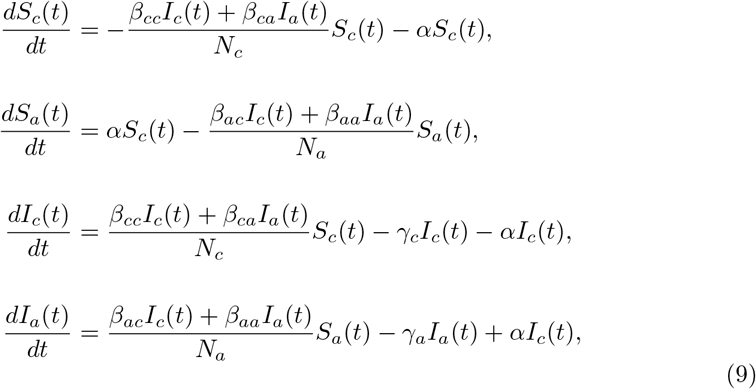

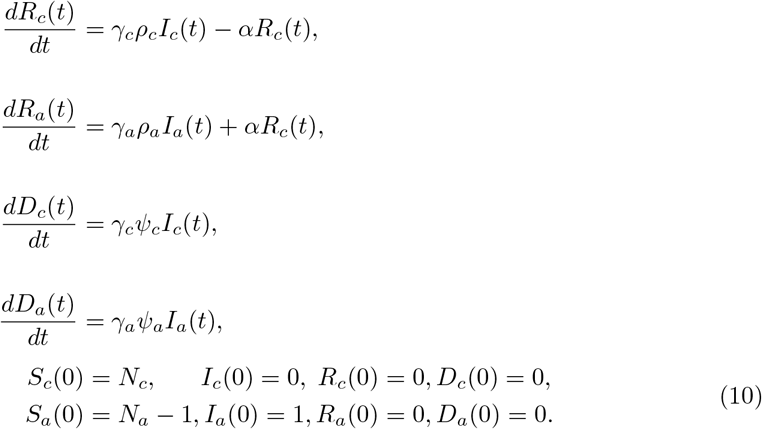

The parameters used in the calculation of the model throughout the paper are presented in Table. 1. The values are cited from the sources themselves except *A* and *β*_*ca*_ calculated from the data of the correlated source [6, 17]. The threshold of the children’s age to become adults in parameter *A* is set to 13 year as the mean value of the group of ages in which the percentage of critical cases relative to all cases is the highest as reported by Dong et al. [6] (Table 2). The susceptible contacts in children who become infected due to direct disease transmission from an adult *β*_*ca*_ are calculated based on data of 10 children. Eight children out of the 10 have been exposed to 30 adults in total and later found to be infected, as reported by Cai et al. [17]. Therefore, the infected rate is set to 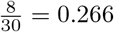.

**Table 1.** Model’s parameters’ description, values, and sources.

| Parameter Definition | Symbol | Value | Source |
| --- | --- | --- | --- |
| Children COVID-19 threshold age [day] | $A$ | 4745 (13 years) | [6] |
| Children to adult conversion rate [day<sup>-1</sup>] | $\alpha := \frac{1}{A}$ | $2.1 \cdot 10^{-4}$ | [6] |
| Infected to recover average duration for children [day<sup>-1</sup>] | $\gamma_c$ | 0.5 | [6] |
| Infected to recover average duration for adults [day<sup>-1</sup>] | $\gamma_a$ | 0.0714 | [8] |
| Susceptible contacts in children which become infected due to direct disease transmission from an adult [day<sup>-1</sup>] | $\beta_{ca}$ | 0.266 | [17] |
| Susceptible contacts in adults which become infected due to direct disease transmission from children [day<sup>-1</sup>] | $\beta_{ac}$ | $\sim 0$ | [19] |
| Susceptible contacts in children which become infected due to direct disease transmission from children [day<sup>-1</sup>] | $\beta_{cc}$ | 0.308 | [18] |
| Susceptible contacts in adults which become infected due to direct disease transmission from an adult [day<sup>-1</sup>] | $\beta_{aa}$ | 0.308 | [18] |
| The rate an infected adult will recover from the disease [day<sup>-1</sup>] | $\rho_a$ | 0.942, 0.98 | [28, 29] |
| The rate an infected child will recover from the disease [day<sup>-1</sup>] | $\rho_c$ | $\sim 1$ | [30] |
| The rate an infected adult will not recover from the disease [day<sup>-1</sup>] | $\psi_a$ | 0.02, 0.05 | [19, 29] |
| The rate an infected child will not recover from the disease [day<sup>-1</sup>] | $\psi_c$ | $\sim 0$ | [30] |

**Table 2.** Markov chain transformation matrix representation of Eq (1-8).

| | $S_c$ | $S_a$ | $I_c$ | $I_a$ | $R_c$ | $R_a$ | $D_c$ | $D_a$ |
| --- | --- | --- | --- | --- | --- | --- | --- | --- |
| $S_c$ | $1 - \alpha - v_1(t)$ | $\alpha$ | $v_1(t)$ | 0 | 0 | 0 | 0 | 0 |
| $S_a$ | 0 | $1 - v_2(t)$ | 0 | $v_2(t)$ | 0 | 0 | 0 | 0 |
| $I_c$ | 0 | 0 | $1 - \alpha - \gamma_c$ | $\alpha$ | $\gamma_c \rho_c$ | 0 | $\gamma_c \psi_c$ | 0 |
| $I_a$ | 0 | 0 | 0 | $1 - \gamma_a$ | 0 | $\gamma_a \rho_a$ | 0 | $\gamma_a \psi_a$ |
| $R_c$ | 0 | 0 | 0 | 0 | $1 - \alpha$ | $\alpha$ | 0 | 0 |
| $R_a$ | 0 | 0 | 0 | 0 | 0 | 1 | 0 | 0 |
| $D_c$ | 0 | 0 | 0 | 0 | 0 | 0 | 1 | 0 |
| $D_a$ | 0 | 0 | 0 | 0 | 0 | 0 | 0 | 1 |

### Numerical solution

To obtain a better understanding of how different parameter values influence the system dynamics, we illustrate the behavior of the system using numerical analysis. Eq (1-8) are ODEs, first-order, nonlinear, from ℝ to ℝ^8^, where ℝ is time (marked by *t*) and ℝ^8^ is the population distribution of all eight populations (marked by *S*_*c*_(*t*), *S*_*a*_(*t*), *I*_*c*_(*t*), *I*_*a*_(*t*), *R*_*c*_(*t*), *R*_*a*_(*t*), *D*_*c*_(*t*), and *D*_*a*_(*t*)). We processed eight-order Runge–Kutta integration on the differential system to enable numerical simulations. Runge–Kutta integration of the equations was implemented by *Octave* programming (version 5.2.0), using standard program *lsode*, for a set of initial conditions described in Eq (10).

The solutions of the system (9-10) are shown in Fig 3. In addition, the population size, adults, and children are set to be *N* = 8000000, *N*_*c*_ = 2240000, *N*_*a*_ = 5760000 to present the distribution of the Israeli population in 2017 as published by the Israeli central bureau of statistics. Fig 3 shows the population group sizes of *S*_*c*_(*t*), *S*_*a*_(*t*), *I*_*c*_(*t*), *I*_*a*_(*t*), *R*_*c*_(*t*), *R*_*a*_(*t*), *D*_*c*_(*t*), and *D*_*a*_(*t*) where the x-axis in all eight graphs represents the time (in days) that has passed from the beginning of the dynamics and the y-axis is the size of each population, respectively. A maximum in the percent of infected children and adults (39%, 89%) is reached in the 8th and 11th days as shown in Fig 3. Therefore, the maximum infected population was 89% of the whole population. Besides, all children infected and recovered after 17 days while all adults recovered after 82 days.

**Fig 3.**
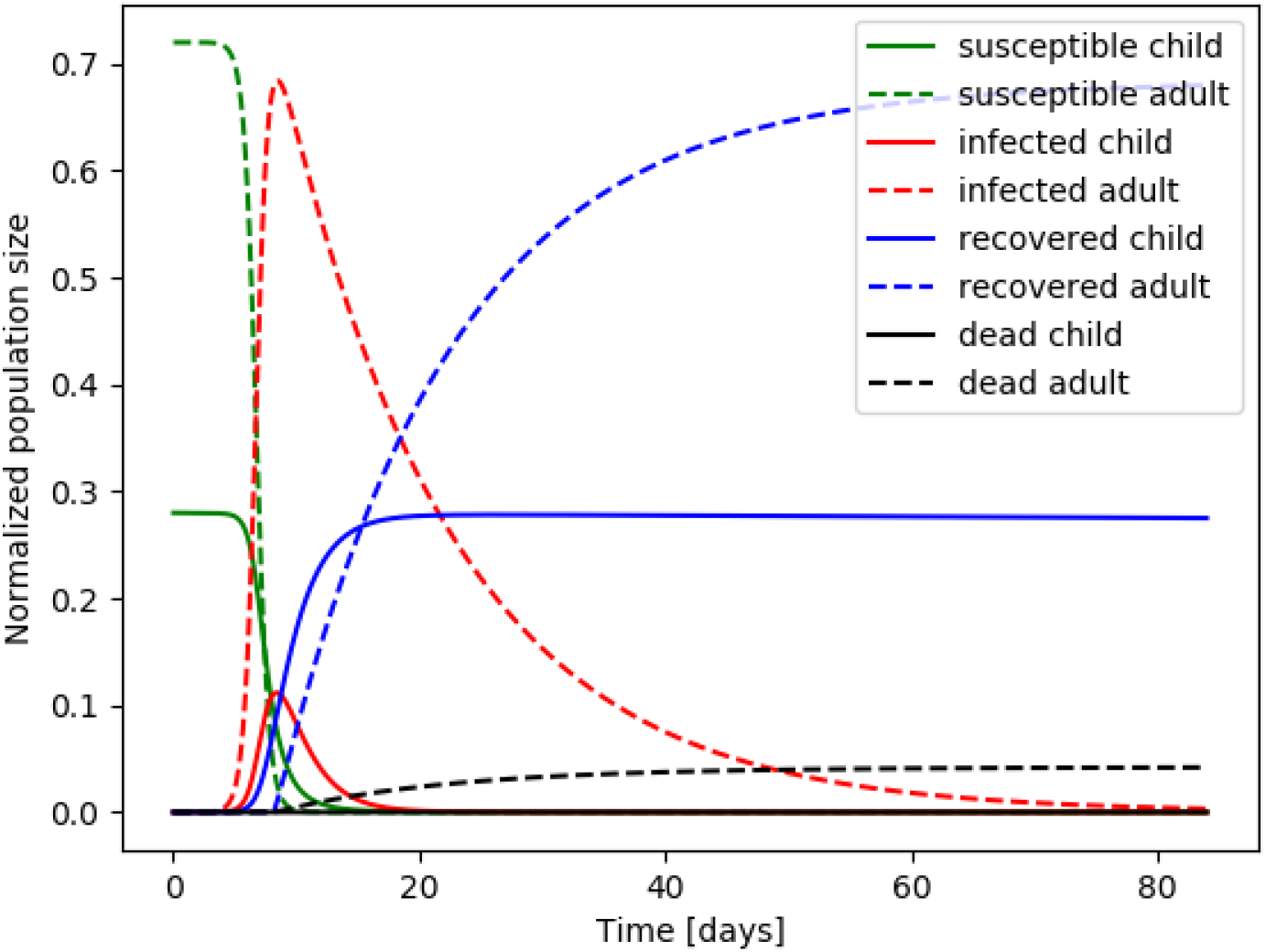
Numerical simulation of trajectories of Eq (9 - 10) and the parameter values from Table. 1. The graphs show the evolution in time (days) of *S*_*c*_(*t*), *S*_*a*_(*t*), *I*_*c*_(*t*), *I*_*a*_(*t*), *R*_*c*_(*t*), *R*_*a*_(*t*), *D*_*c*_(*t*), and *D*_*a*_(*t*). Adult and children graphs are presented with a dotted and solid lines, respectively. Susceptible, infected, recovered, and dead are shown in green, red, blue, and black, respectively.

### Stability analysis

In epidemiology, it is essential to quantify the severity of outbreaks of infectious diseases. The standard parameter indicates the severity called *the basic reproduction number R*_0_. In the standard SIR model [22], *R*_0_ is defined to be the ratio between individual infection rate and recovery rate, where almost everyone is susceptible (namely, *S*_*c*_ + *S*_*a*_ *∼ N*). For any start condition and parameters, at *t →∞* the model’s state is an instance of Eq (10) according to Lemma 1.

#### Definition 1.

The model’s state at time *t* is defined by

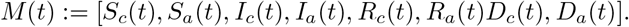

#### Lemma 1.

Assume *α, γ*_*a*_, *γ*_*c*_, *β*_*aa*_, *β*_*cc*_ *>* 0. The asymptotic state *M* (*t*) of the model takes the form:

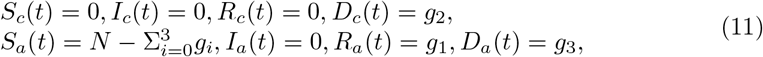

where *g* is a function *g*: ℝ^17^ *→*ℝ^3^ = [*g*_1_, *g*_2_, *g*_3_] such that *g*(ℙ), where

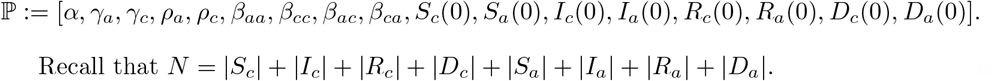

Recall that N = |*S*_*c*_| + |*I*_*c*_ | + |*R*_*c*_ | + |*D*_*c*_ | + |*S*_*a*_ | + |*I*_*a*_ | + |*R*_*a*_ | + |*D*_*a*_ |.

*Proof*. Define a non-homogeneous continuous-time Markov chain *M* with state space with eight values in respect to *S*_*c*_, *S*_*a*_, *I*_*c*_, *I*_*a*_, *R*_*c*_, *R*_*a*_, *D*_*c*_, and *D*_*a*_. Table 2 represents the transition matrix of *M* where

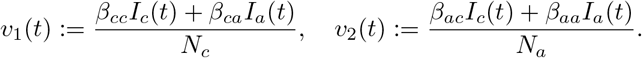

In order to model the dynamics as a Markov chain one needs to show that

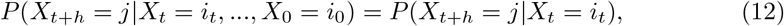

where *{X}*_*t*_ is a stochastic process with values in the state space for all *t ≥* 0 and all states *i*_0_, …*i*_*t*_, *j* and *h* is an arbitrary small step in time [31]. *M* satisfies Eq (12) if both *v*_1_(*t*), *v*_2_(*t*) depend only on the last state *X*_*t*_ = *i*_*t*_.

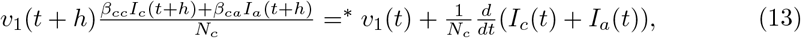

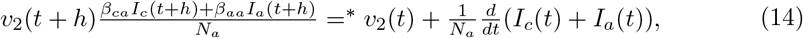

such that (=^*∗*^) stands for *I*_*c*_(*t*+*h*) = *I*_*c*_(*t*)+*dI*_*c*_(*t*), *I*_*a*_(*t*+*h*) = *I*_*a*_(*t*)+*dI*_*a*_(*t*). From Eq (13) and (14), *v*_1_(*t* + *h*) and *v*_2_(*t* + *h*) depends only on the previous state *X*_*t*_ = *i*_*t*_, respectively. Let *π* ∈ ℕ^8^ be the initial condition for the system. Therefore, the model’s state at some time *t* is defined by *A*^*t*^*π*. For any initial condition *π*, the vector lim_*t→∞*_(*A*^*t*^*π*) takes the form

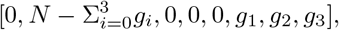

where *g*_1_, *g*_2_, *g*_3_ ∈ [0, …, *N*].

From Table 2 one can see that *R*_*a*_, *D*_*c*_, and *D*_*a*_ are stationary values because 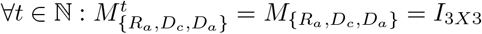. By setting *π* = [*N*_*c*_, *N*_*a*_, 0, 0, 0, 0, 0, 0] one gets *∀t* ∈ℕ: *v*_1_(*t*) = *v*_2_(*t*) = 0 and therefore the sub matrix of *M* with only the states *S*_*c*_ and *S*_*a*_ takes the form

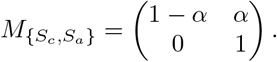

Therefore, *M* (*t → ∞*) = [0, *N*, 0, 0, 0, 0, 0, 0] because

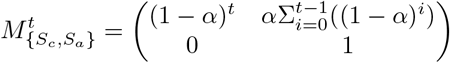

and for *lim*_*t→∞*_(1 *− α*)^*t*^ = 0 meaning that asymptotically the population in *S*_*c*_ decreases to zero. Therefore, the asymptotic state of the model takes the form:

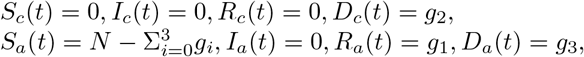

for any initial condition *π* and parameters such that *α, γ*_*a*_, *γ*_*c*_, *β*_*aa*_, *β*_*cc*_ *>* 0.

In order to find *g* (from Lemma 1) for given initial conditions and parameters, it is possible to use the next generation matrix approach [32]. Let 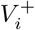 be the rate of transfer of individuals by all means except of appearance of new infections in compartment *i*, and 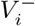 be the rate of transfer of individuals out of compartment *i*. Let *F*_*i*_ be the rate of appearance of new infections in compartment *i*. Eq (1 - 8) takes the form:

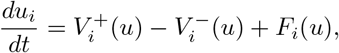

where

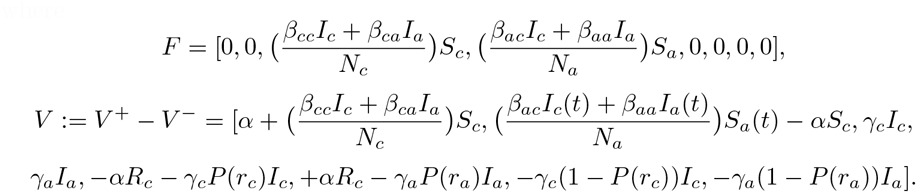

In equilibrium, *I*_*c*_ and *I*_*a*_ are both equal to zero so the derivatives at equilibrium, focusing on *I*_*c*_ and *I*_*a*_ from Eqs. (3) and (4), are mapped to the third and forth elements in vectors *F* and *V* giving the matrices **F** and **V**.

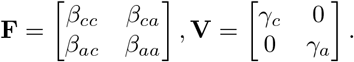

The next generation matrix is defined as **FV**^*−*1^ [32].

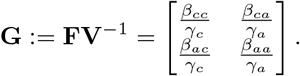

Assume an initial condition

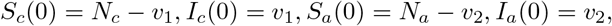

marked as *v* = [*v*_1_, *v*_2_]. After one time unit, the amount of infected individuals can be calculated using *v*^*′*^ = **G***v*. Therefore, for any start condition and model parameters one can find *g* by calculating

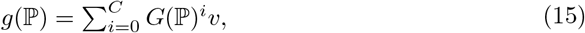

where *C* ∈ℕ is the first index that satisfies *I*_*c*_(*C*) + *I*_*a*_(*C*) = 0.

It is possible to retrieve *R*_0_ from the next generation matrix G as it is the dominant eigenvalue of the matrix [25], as this matrix describes the total amount of infections caused by each class over the lifetime of the infection. The epidemic is assumed to be stable if *R*_0_ *≤* 1. This means that for each infected individual there is less than one infected individual in any of the groups in the next time unit. The dominant eigenvalue of **G** can be calculated using the characteristic equation which is a second-order polynomial of *R*_0_. The zero points of this polynomial are

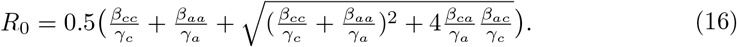

Both *γ*_*c*_ and *γ*_*a*_ are biological properties of the disease; on the other hand, *β*_*aa*_, *β*_*ac*_, *β*_*ca*_, and *β*_*cc*_ can be managed using social distance, quarantine, masks, and other methods. Figs 4a - 4e presents five projections of stability from the parameter space *{β*_*aa*_, *β*_*ac*_, *β*_*ca*_, *β*_*cc*_*}* calculated using Eq (16). Fig 4a shows the *β*_*aa*_ *− β*_*ac*_ projection where *β*_*cc*_ = 0.308, *β*_*ca*_ = 0.266. There are no values such that *R*_0_ *<* 1 and from the color gradient, it is possible to see that *β*_*ac*_ has slightly more influence on *R*_0_ relatively to *β*_*aa*_. Fig 4b shows the *β*_*aa*_ *− β*_*ca*_ projection where *β*_*cc*_ = 0.308, *β*_*ac*_ = 0. There are no values such that *R*_0_ *<* 1 and from the color gradient, it is possible to see that *β*_*aa*_ has a minor to no influence on *R*_0_ relatively to *β*_*ca*_. Fig 4c shows the *β*_*cc*_ *− β*_*ca*_ projection where *β*_*aa*_ = 0.308, *β*_*ca*_ = 0.266. *R*_0_ *<* 1 for any combination of *β*_*cc*_, *β*_*ac*_ such that *β*_*ac*_ *<* 0.08 *−* 1.6*β*_*cc*_. Fig 4d shows the *β*_*cc*_ *− β*_*ac*_ projection where *β*_*aa*_ = 0.308, *β*_*ac*_ = 0. *R*_0_ *<* 1 for any combination of *β*_*cc*_, *β*_*ac*_ such that *β*_*cc*_ *≤* 0.3. Fig 4e shows the *β*_*aa*_ *− β*_*cc*_ projection where *β*_*ca*_ = 0.266, *β*_*ac*_ = 0. *R*_0_ *<* 1 for any combination of *β*_*aa*_, *β*_*cc*_ such that *β*_*cc*_ *<* 0.9 *−* 0.155*β*_*aa*_.

**Fig 4.**
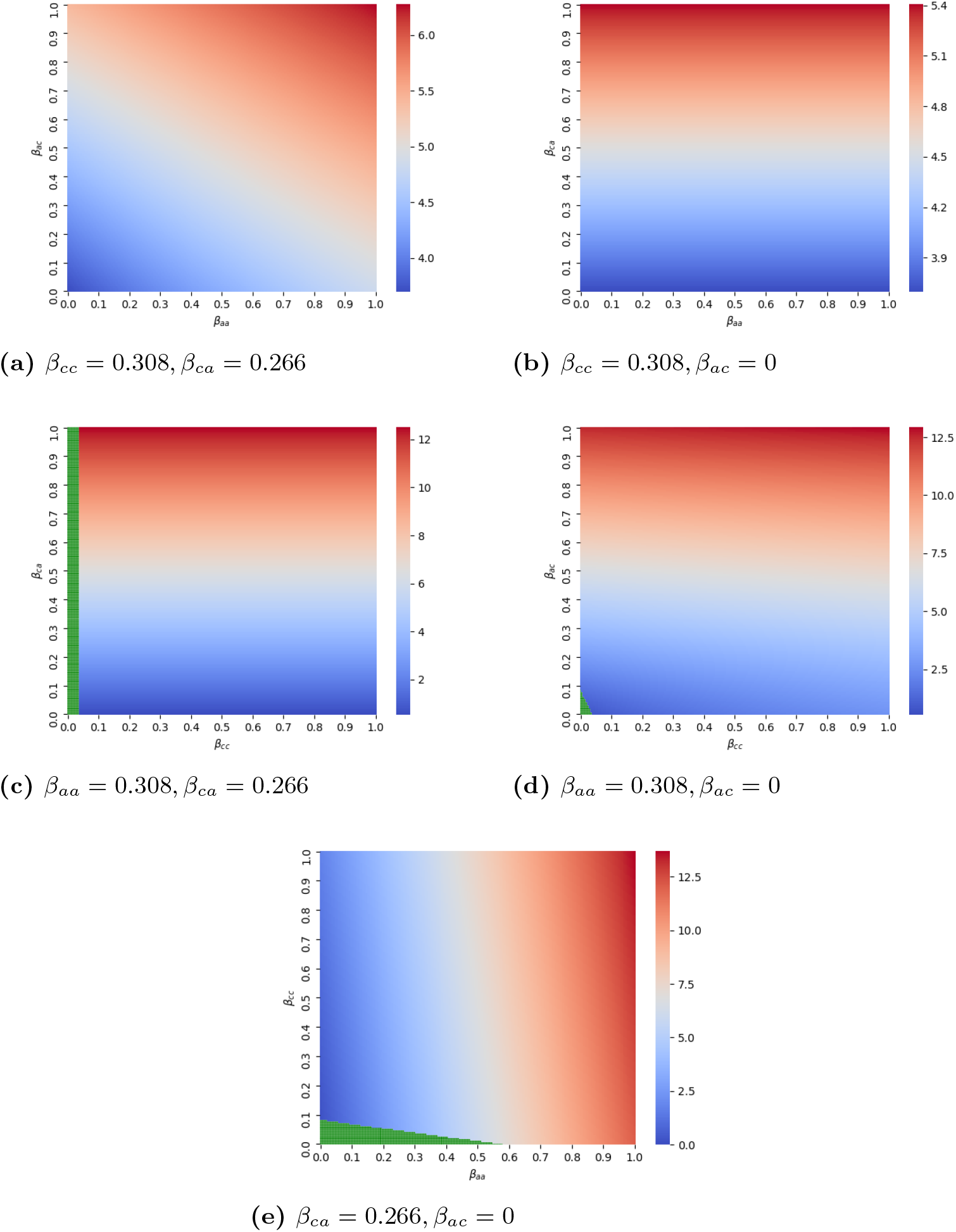
2-dimensional projections of the *{β*_*aa*_, *β*_*ac*_, *β*_*ca*_, *β*_*cc*_*}* space and there influence on *R*_0_. The green section is where *R*_0_ *≤* 1. The color gradient is from blue (lower *R*_0_) to red (higher *R*_0_). The model parameters used are *α* = 8.78 *·* 10^*−*6^, *γ*_*c*_ = 0.5, *γ*_*a*_ = 0.0714.

In order to obtain the equilibria points of the model and their stability properties, the Jacobian (*J*) is obtained by linearizing Eqs (3) and (4) in the system (Eqs (1 - 8)). The calculations show that

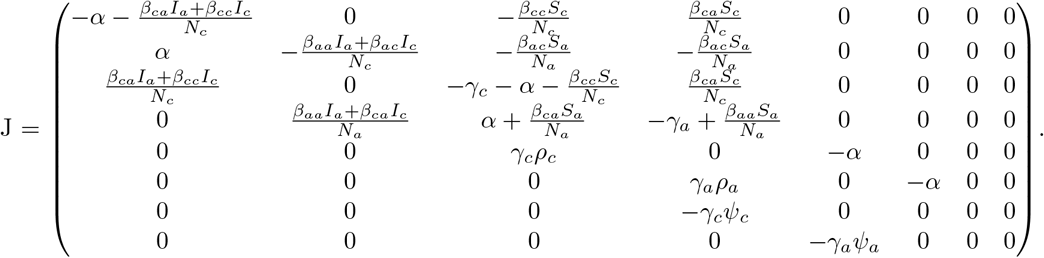

The system (Eqs (9-10)) has four non-trivial equilibria which may be found by setting all rates in Eq (9) to zero; marked by *EQ*_1_, *EQ*_2_, *EQ*_3_, and *EQ*_4_. Thus equilibria are provided as the state vector of the eight population states in Table 3, where

**Table 3.** The four equilibria solutions for Eqs (9-10).

| | $EQ_1$ | $EQ_2$ | $EQ_3$ | $EQ_4$ |
| --- | --- | --- | --- | --- |
| $S_c$ | 0 | $\frac{(\gamma_c - \alpha)N_c}{\beta_{cc}}$ | $\frac{\beta_{ca}N_a^2(\alpha - \gamma_a)}{\beta_{aa}N_c}$ | $\frac{(\alpha + \gamma_c)I_c^*}{\beta_{cc}I_c + \beta_{ca}I_a}$ |
| $S_a$ | N | $\frac{\beta_{ca}N_c^2(\gamma_c - \alpha)}{N_a\beta_{cc}}$ | $\frac{(\alpha - \gamma_a)N_a}{\beta_{aa}}$ | $\frac{(\gamma_a - \alpha)I_c^*}{\beta_{ac}I_c^* + \beta_{aa}I_a^*}$ |
| $I_c$ | 0 | $\frac{\alpha N_c}{\beta_{cc}}$ | $\frac{\alpha N_c}{\beta_{ca}}$ | $I_c^*$ |
| $I_a$ | 0 | 0 | $\frac{\gamma_a \rho_a N_c}{\beta_{ca}}$ | $I_a^*$ |
| $R_c$ | 0 | $\frac{\gamma_c \rho_c N_c}{\beta_{cc}}$ | 0 | $\frac{\gamma_c \rho_c I_c^*}{\alpha}$ |
| $R_a$ | 0 | 0 | $\frac{\alpha N_c}{\beta_{ca}}$ | $\frac{-\gamma_a \rho_a I_a^*}{\alpha}$ |
| $D_c$ | 0 | $\frac{\alpha N_c}{\beta_{cc}} - \frac{-\gamma_c \rho_c N_c}{\beta_{cc}}$ | 0 | $I_c^* - \frac{\gamma_c \rho_c I_c^*}{\alpha}$ |
| $D_a$ | 0 | 0 | $\frac{\alpha N_c}{\beta_{ca}} - \frac{\gamma_a \rho_a N_c}{\beta_{ca}}$ | $I_a^* + \frac{\gamma_a \rho_a I_a^*}{\alpha}$ |

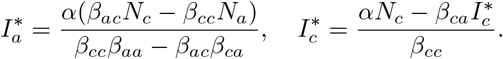

There are other equilibria for trivial cases such as *S*_*a*_(0) = 0 or *S*_*c*_(0) = 0 which are unrealistic for real-world dynamics. Below we investigate the stability of four nontrivial equilibria.

*EQ*_1_: is the case for *I*_*c*_(*t*) = *I*_*a*_(*t*) = 0 for every *t* ∈ℕ (see Table 3). This is a trivial equilibrium where there is no epidemic at all. Because the model does not take into consideration the birth of new children, after *A* days all children become adults and *EQ*_1_ is derived. By settings *J* (*EQ*_1_), all eigenvalues are negative for all parameter values except 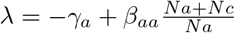. Therefore, this equilibrium is stable if 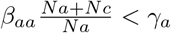.

*EQ*_2_: is the case for *I*_*a*_(*t*) = 0 for every *t* ∈ℕ (see Table 3). This equilibrium can be achieved if *β*_*ac*_ = 0 because in any other case (assuming *S*_*a*_(0) *>* 0) exists time *t* such that 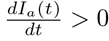 and therefore *I*_*a*_(*t* + 1) ≠ 0.

*EQ*_3_: is the case for *I*_*c*_(*t*) = 0 for every *t* ∈ℕ (see Table 3). This equilibrium can be achieved if *β*_*ca*_ = 0 because in any other case (assuming *S*_*c*_(0) *>* 0) exists time *t* such that 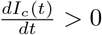 and therefore *I*_*c*_(*t* + 1) ≠ 0.

*EQ*_4_: is the case for ∃*t* such that *I*_*c*_(*t*) ≠ 0 and *I*_*a*_(*t*) ≠ 0 (see Table 3). This equilibrium is the generic case for the model.

According to Lemma 1, *EQ*_2_, *EQ*_3_, and *EQ*_4_ are not possible asymptotic forms of the system and are therefore not stable.

### Spatial model

Fig 5 shows the spatial model schema presenting the populations distribution and locations (work / school, and home) at some time of the day. In addition to the ODE model’s parameters (shown in Table 1), the follows parameters are added to the *hybrid* model as part of the spatial model:

**Fig 5.**
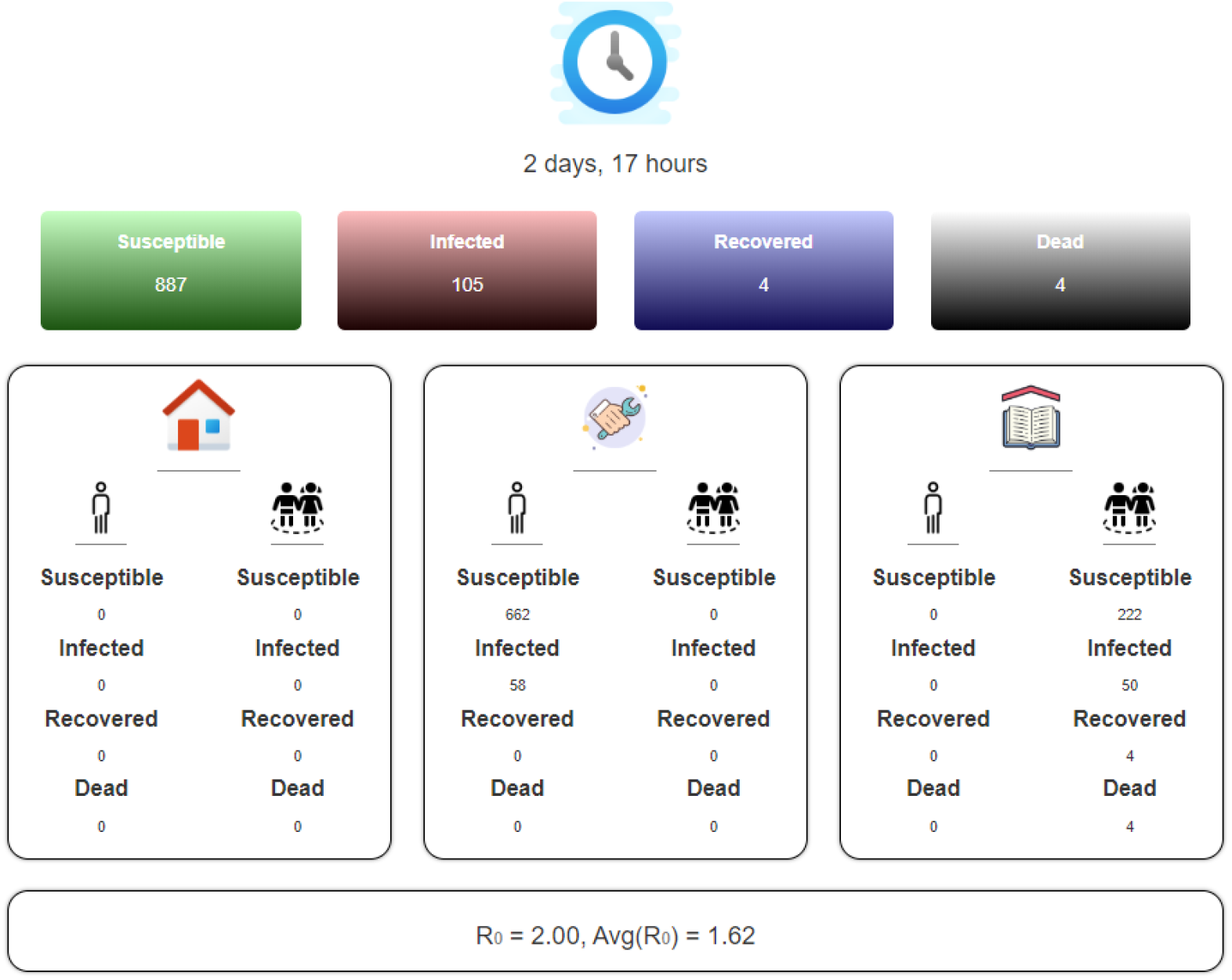
The panel of the spatial model. From top to bottom: the time from the beginning of the simulation. Distribution of the population to susceptible, infected, recovered, and dead groups. The distribution of the population to susceptible, infected, recovered, and dead groups with separation to children and adults and their current location (home, work, school). The *R*_0_ at a certain time and the average *R*_0_ from the beginning of the simulation.

1. *ϕ*_*ac*_ the average number of meeting events between adults and children per hour.
2. *ϕ*_*aa*_ the average number of meeting events between adults and adults per hour.
3. *ϕ*_*cc*_ the average number of meeting events between children and children per hour.
4. 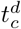 hours of the day that children are at home and 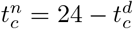 hours of the day that children are at school.
5. 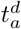 hours of the day that adults are at home and 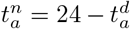 hours of the day that adults are at work.

We assume the transition from home to either work or school and back is immediate and that everybody is following the same clock. Each simulation step simulates one hour. The population size is constant during the simulation and initialized in the beginning of each iteration by setting children population size *N*_*c*_ and adult population size *N*_*a*_. In each simulation step, the following three actions take place:

1. If a member is in the susceptible group and meets other members of the infected group there is a change of *β*_*aa*_, *β*_*ac*_, *β*_*ca*_, or *β*_*cc*_ according to the age-class of the two members that the first will be infected.
2. Each infected child or adult that was infected for 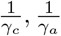 simulation steps becomes either recovered or deceased, respectively.
3. According to the hour of the day, the adults transition to home or work and the children to home or school.

The spatial model adds day-night circle and three main locations to the dynamics of the *hybrid* model.

Fig 6 shows the population group sizes of *S*_*c*_(*t*), *S*_*a*_(*t*), *I*_*c*_(*t*), *I*_*a*_(*t*), *R*_*c*_(*t*), *R*_*a*_(*t*), *D*_*c*_(*t*), and *D*_*a*_(*t*) as an average of 10 iterations of the *hybrid* model. The x-axis is the time (in days) that has passed from the beginning of the epidemic and the y-axis is the normalized size of each population, respectively. The parameters used in the simulations are 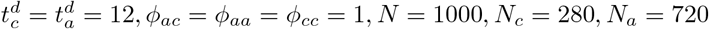.

**Fig 6.**
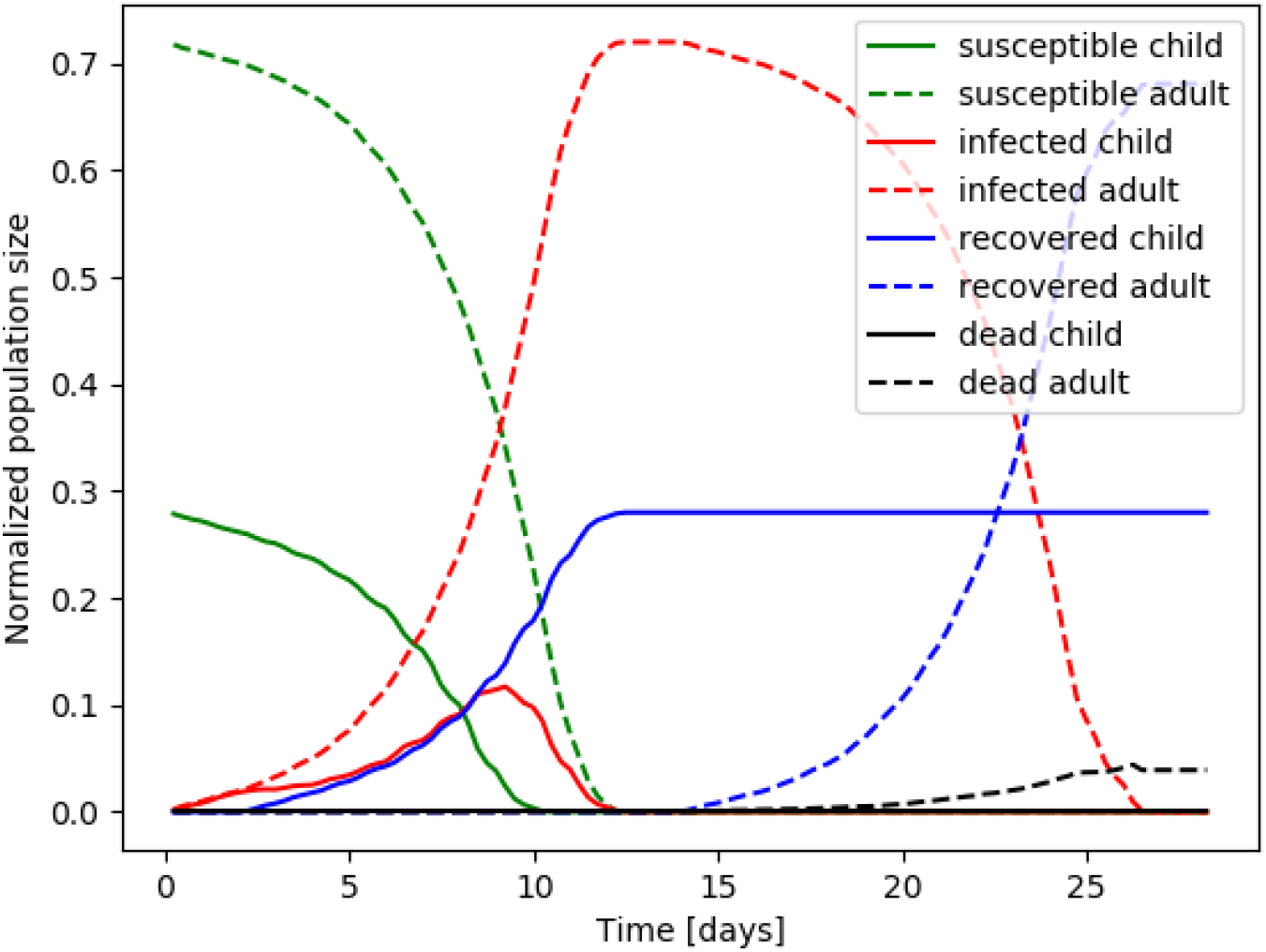
Average of 10 iteration of numerical simulation of the *hybrid* model where the parameter values are taken from Table. 1. Children and adult graphs are presented with a dotted and solid lines, respectively. Susceptible, infected, recovered, and dead are shown in green, red, blue, and black, respectively.

A maximum in the percent of infected children and adults (38.5%, 100%) is reached on the 9_*th*_ and 12_*th*_ day, as shown in Fig 6. In addition, all children were infected and recovered after 12 days while all adults recovered after 28 days. The simulation shows similar results to the results derived by solving the model (Eqs (9-10)) as presented in Fig 3. The main difference between the two is that the simulation predicts 4.43 times shorter duration from the time of maximum infected adults to the time that all adults are either dead or recovered, as the infected adult population equals zero on the 82_*nd*_ and 28_*th*_ days. In addition, the maximum of infected adults is reached on the 11_*th*_ and 12_*th*_ days resulting in 71 and 16 days for full recovery.

## Results

### Outbreak analysis

The proposed *hybrid* model provide an *in-silico* environment allowing relatively fast, cheap, and accurate analysis of different policies on the COVID-19 spread dynamics.

One of the major hopes of politicians is for a NPI policy in which the epidemic does not reach an outbreak at any time after they initialized a given decision. We define this condition mathematically as follows:

#### Definition 2.

For given initial condition and model’s parameter (ℙ). A solution of Eq (9-10) is defined as *outbreak* dynamics if ∃*t* ∈ ℕ: *R*_0_(*t*) *>* 1.

We will examine two policies, based on this condition, to determine if each one is possible to fulfill the condition. If so, the optimal NPI policy is based on the parameter-space criteria:

1. The influence of the duration of the work/school day.
2. Lockdown in homes with partial to full separation between individuals.

### Duration of working and schooling day

We will assume that children meet only children in school and adults meet only adults at work and at home adults and children meet each other. This is a good approximation of the real dynamics as a relatively small percent of adults work with children during the day which keeps the interactions between children and adults at this time relatively small to the extent of interaction when both adults and children are at home and therefore can be neglected.

Based on the proposed spatial model, we change the number of hours children and adults spend in school and work each day, respectively. Fig 7a shows the average *R*_0_ as a function of the duration in hours of the working (*d*_*W*_) and schooling (*d*_*S*_) day. The dots are the calculated values from the simulator and the surface is a fitting function *R*_0_(*d*_*W*_, *d*_*S*_). The fitting function is calculated using the least mean square (LMS) method [33]. In order to use the LMS method, one needs to define the family function approximating a function. The family function

**Fig 7.**
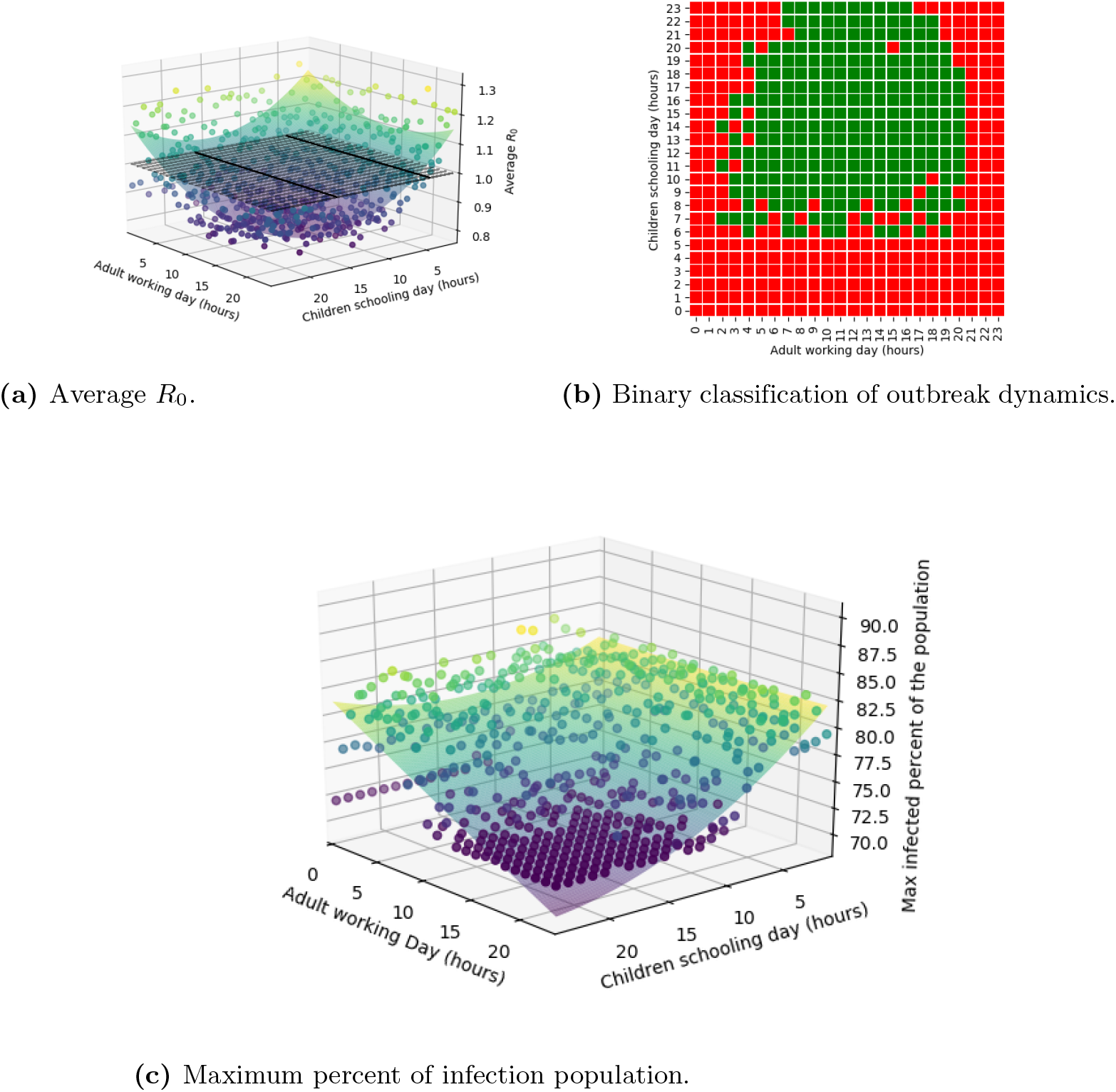
Analysis of the epidemic spread as a function of working (*d*_*W*_) and schooling (*d*_*S*_) duration in hours each day.

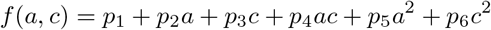

has been chosen to balance between the accuracy of the sampled data on the one hand and simplicity of usage on the other [34],

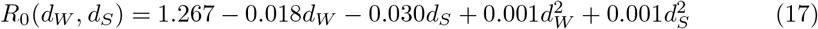

and was obtained with a coefficient of determination *R*^2^ = 0.815. Similarly, Fig 7c shows the average max_*t*_(*I*_*c*_(*t*) + *I*_*a*_(*t*)) as a function of the duration in hours of the working (*d*_*W*_) and schooling (*d*_*S*_) day. The dots in Fig 7c are the calculated values from the spatial model and the surface is the fitting function

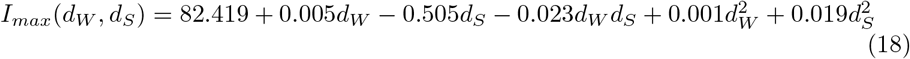

obtained with a coefficient of determination *R*^2^ = 0.748.

The duration of either working or schooling day has a local minimum at (*d*_*W*_, *d*_*S*_) = (18, 14) with *R*_0_ = 0.808, as shown in Fig 7a. The optimal point from Eq (17) is (*d*_*W*_, *d*_*S*_) = (9, 17.5) with *R*_0_ = 0.967 obtained by setting the gradient of *R*_0_(*d*_*W*_, *d*_*S*_) to zero. The inconsistency in the values is resulted by the error in the fitting function but both show the same behavior in which longer working hours reduce significantly the infection rate but too many hours increase the infection rate back as the lack of circulation of contagious adults and children during the day helps to reduce the infection rate. Furthermore, Fig 7b presents a binary classification where red cells are cases with an outbreak and green cells are cases without outbreak during the simulation as a function of the working (*d*_*W*_) and schooling (*d*_*S*_) duration in hours each day.

On the other hand, the duration of either working or schooling day has an effect of 10% (as the maximal value is 82.5 while the minimal is 72.5) on the maximal infected percent of individuals from the population as shown in Fig 7c and by setting the limits *d*_*W*_ = {0, 24}, *d*_*S*_ = 0, 24 in Eq (18). There is a sharp decline between a shorter working-schooling duration of 10 hours or less and longer than that. This is associated with the fact that with more than 12 hours (half-day) the dynamics that have a higher incidence are the ones with *β*_*ca*_ = *β*_*ac*_ = 0 which reduces the number of infected individuals in total.

### Lockdown policy

Partial or full lockdown of several locations or entire countries were broadly used at the beginning of the COVID-19 outbreak as a policy to reduce the number of infected individuals and control the spread dynamics. The lockdown policy yields social distance which reduces the ratio of infection. On the other hand, this policy negatively affects the economy and mental health, and increases the presence of other diseases (both physical and mental) in the population. Therefore, the optimization task of finding the minimal portion of the population to be locked down such that the epidemic will be constrained is important.

The optimization problem can be written formally as

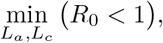

where *L*_*a*_ and *L*_*c*_ are the portion of adults and children in lockdown. We ran the spatial model multiple times where each time a combination of 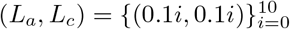 of the population is assumed to be in lockdown. Fig 8a shows the results of this calculation. Each dot represents *R*_0_ of each case *L*_*a*_, *L*_*c*_. The black grid shows the threshold *R*_0_ = 1.

**Fig 8.**
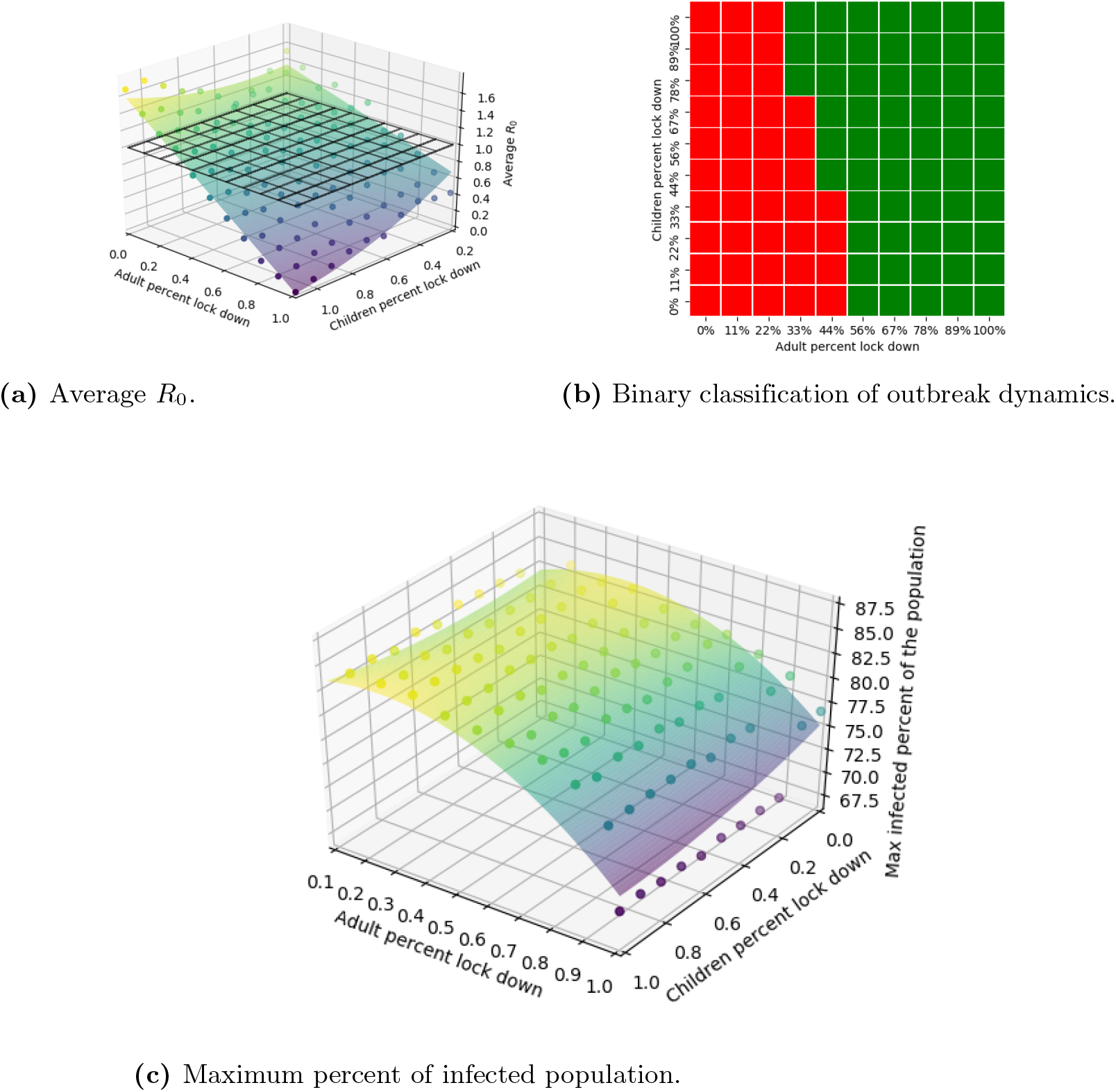
Analysis of the epidemic spread as a function of adult lockdown (*L*_*a*_) and children lockdown (*L*_*c*_) using the computer simulation of the *hybrid* model.

The behavior of *R*_0_ as a function of *L*_*a*_, *L*_*c*_ has been retrieved using the LMS method and takes the form

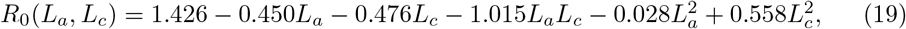

obtained with a coefficient of determination *R*^2^ = 0.971. Therefore, it is safe to claim that function *R*_0_(*a, c*) is well fitting the data despite the stochastic noise of the simulation and presents a fair approximation for the *R*_0_ behavior as a function of *L*_*a*_, *L*_*c*_. Using Eq (19) it is possible to find the constraints of *L*_*a*_, *L*_*c*_ such that *R*_0_ *≤* 1, by solving *R*_0_(*L*_*a*_, *L*_*c*_) *≤* 1.

Closing only schools without any reduction in adults going to work does not prevent an epidemic outbreak while locking down half of the adult population will prevent an outbreak. Lockdown of children (*L*_*c*_) have a minor effect relatively to lockdown adults (*L*_*a*_) as shown in both Eq (19) and Fig 8b. In addition, the same phenomena repeat in the max infected individuals as shown in Fig 8c. The surface calculated using the LMS method:

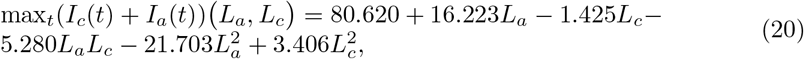

and obtained with a coefficient of determination *R*^2^ = 0.840.

In addition, we repeat the analysis by numerically solving the system (Eq (9-10)). The lockdown is reflected in the model as the infection rate between every two individuals. *L*_*a*_ affects the interactions between adults and either other adults or children *β*_*aa*_, *β*_*ac*_ and *L*_*c*_ affects the interactions between children and either other children or adults *β*_*cc*_, *β*_*ca*_. Mark the original values of *β*_*aa*_, *β*_*ac*_, *β*_*ca*_, *β*_*cc*_ from Table 1 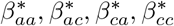 Each time, the system solved where 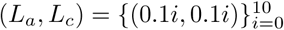 such 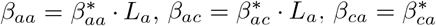 and 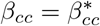.

Fig 9a shows the results of this calculation based on Eq (16). Each dot represents *R*_0_ of each case *L*_*a*_, *L*_*c*_. The black grid shows the threshold *R*_0_ = 1. Fig 9b presents a binary classification of the cases with outbreak dynamics (red) or without (green). Fig 9b is similar to Fig 8b such that adult lockdown has a higher influence on the outbreak and predicts that a much higher percentage of the population will be in lockdown. Again, the difference in the percent of adult’s lockdown (*L*_*a*_) such that *R*_0_ *<* 1 can be associated with the slower infection rate in 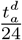 hours of each day because part of the adults change their location which in it turns influence the infection rate. In addition, the recovery process is faster in the spatial model (as shown in Figs 3 and 6).

**Fig 9.**
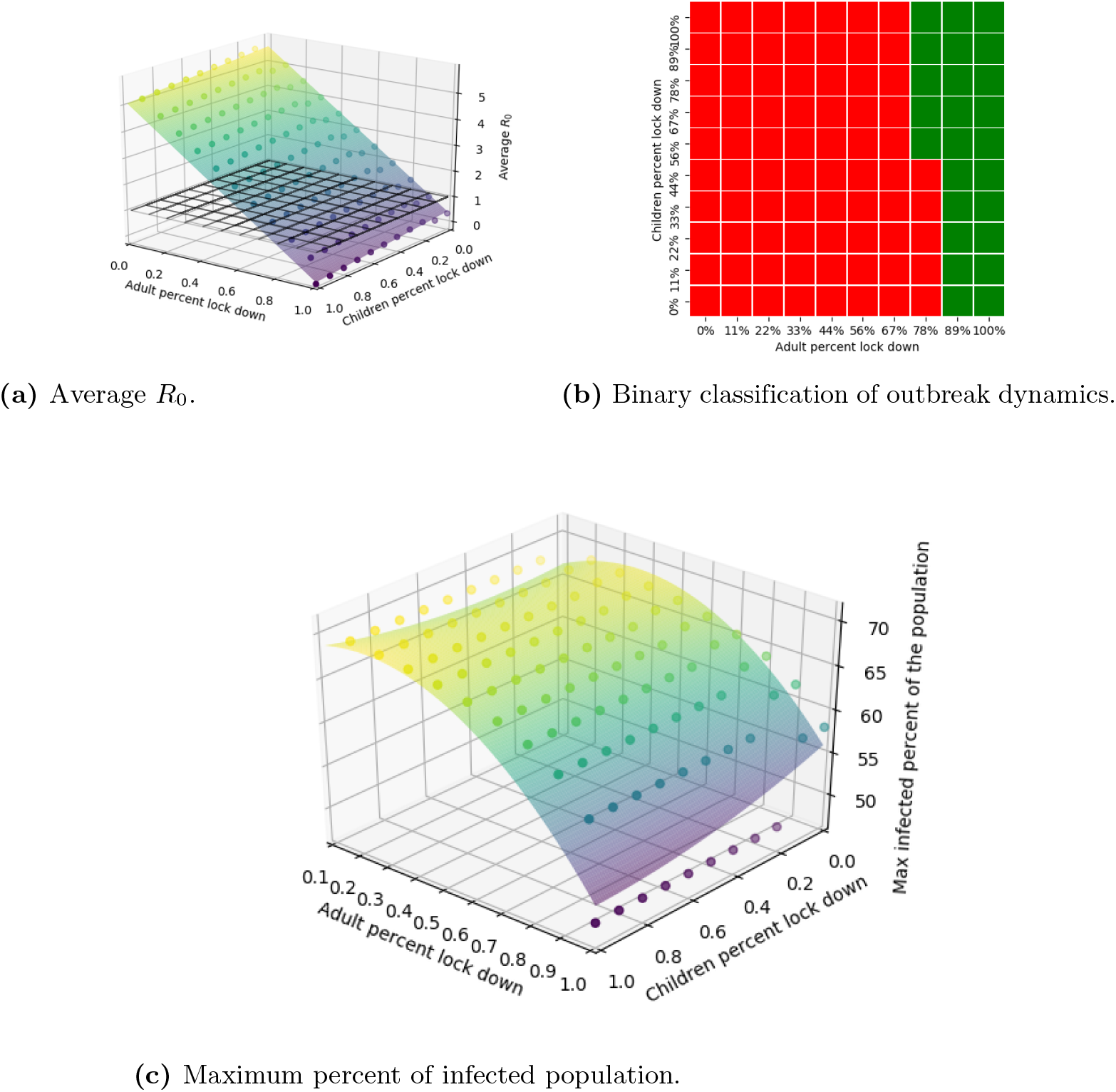
Analysis of the epidemic spread as a function of adult lockdown (*L*_*a*_) and children lockdown (*L*_*c*_) by the *hybrid* model.

The behavior of *R*_0_ as a function of *L*_*a*_, *L*_*c*_ has been derived using the LMS method and takes the form

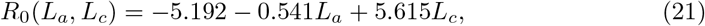

obtained with a coefficient of determination *R*^2^ = 0.952.

In addition, Fig 9c presents the max infected individuals as a function of adult and children lockdown. The surface has been calculated using the LMS method and takes the form:

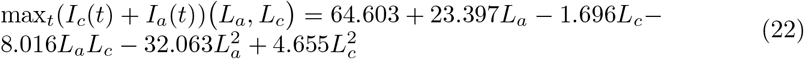

obtained with a coefficient of determination *R*^2^ = 0.702. Figures 8c and 9c are presenting the same behavior while Fig 8c predicts 5% more infected individuals on average. The influence of the adult lockdown is one order of magnitude more significant that of the children’s lockdown in a first and second order approximation as shown in Eqs (20) and (22). That is, where there is no lockdown for children (*L*_*c*_ = 0) there is a local maximum in infected individuals regardless of the lockdown of adults (*L*_*a*_).

### Validation of the *hybrid* model on the dynamics of the COVID-19 pandemic in Israel

In Israel, in February 21 the first COVID-19 infected individual was detected. On March 25 a national quarantine was implemented that continued for two months with local relief and restriction on several occasions and in several cities. Fig 10 presents the daily new confirmed cases from August 15 to September 28, respectively. Where the blue (solid) line marks the date when the schools returned to almost full capacity. The green (solid-dotted) lines are linear regression for before and after school opening, respectively. It is easy to see a significant increment in the number of new daily cases.

**Fig 10.**
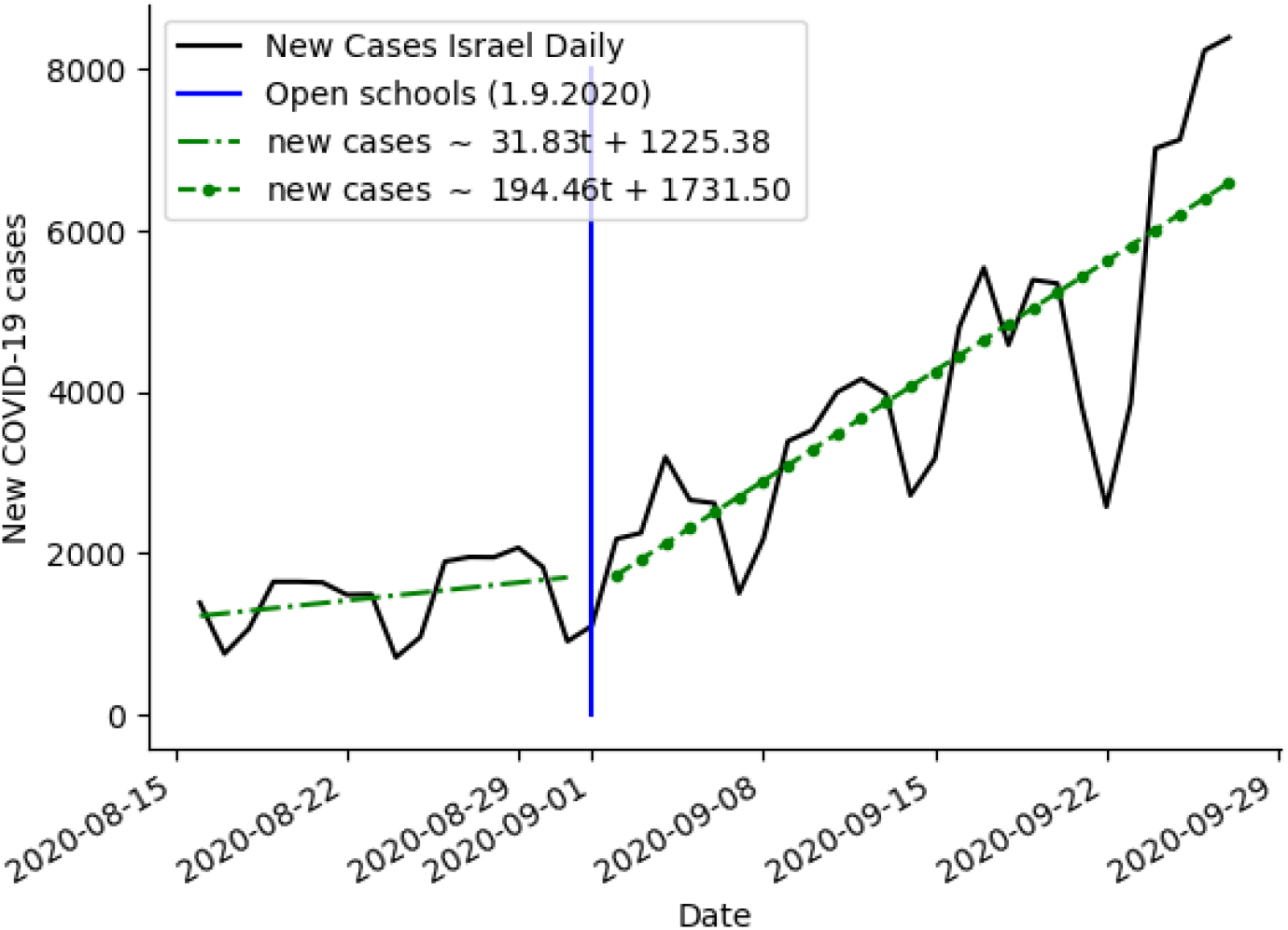
Daily confirmed cases in Israel between August 15 and September 29 (2020) [4]. The blue (solid) line is the full opening of schools. The green (solid-dotted) lines are a linear regression for before and after school opening, respectively.

From [4] there was 90232 infected at August 15. Given that the average recover rate of children is 2 days and adults is 14 days (see Table 1) and that children is 28% of the population, we obtain that there was approximately 18899 infected adults and 868 infected children. We assume that the amount of recovered individuals is heterogeneous in the population and that only adults die from the epidemic until this point. Eq (23) shows the initial conditions for August 15 (*t*_0_), Israel.

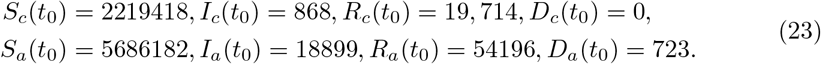

To test the *hybrid* model, we calculated the *R*_0_ on average per day for the period from August 15 to September 1 and from September 1 to September 15, dividing the number of new cases by the total number of cases on that day [4]. We solve numerically the *hybrid* model and calculate *R*_0_ for the case when the schools are open versus the case when the schools are closed. If schools are open, children go to school three times a week, every other day and do not go on weekends for five hours. If schools are closed, children stayed at home all the time. In both cases, we assume that adults will work eight hours each day, except on weekends when they stay home. The obtained values of *R*_0_ for both cases are shown in Fig 11 with mean square error (MSE) of 0.205.

**Fig 11.**
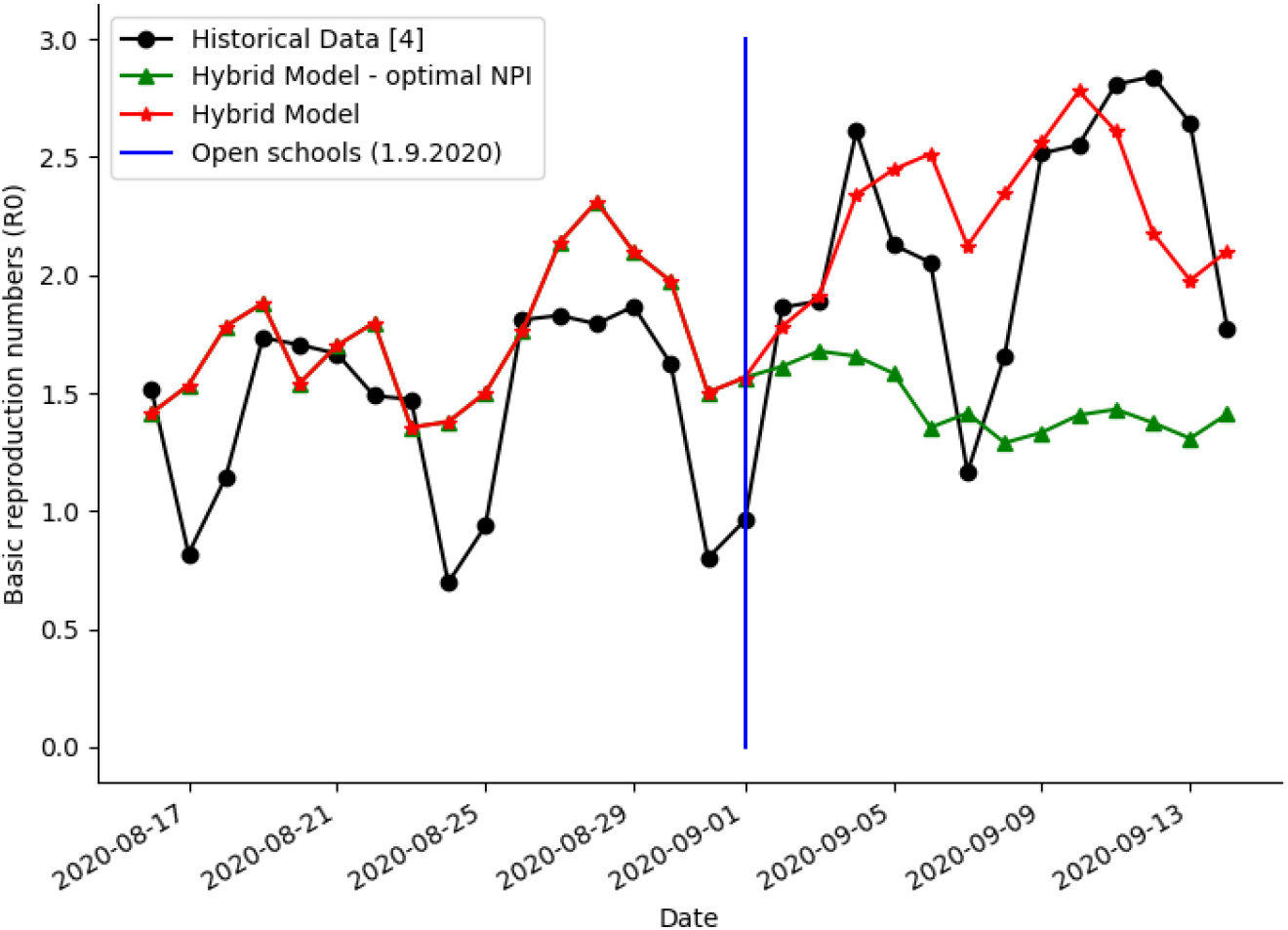
Daily *R*_0_ between August 15 and September 15 (2020) comparison between the historical data and the *hybrid* models predictions.

Considering that, longer school day is reducing the infection rate (Fig 7a), we solve numerically the *hybrid* model and calculate *R*_0_ for the case when the schools are open (from September 1). We assume that children go to school every day (except weekends) for nine hours while the adults go to work for the same period. We obtain that the difference average *R*_0_ of the case with the optimal NPI (*R*_0_ = 1.45) and the historical NPI (*R*_0_ = 2.28) is Δ*R*_0_ = 0.83 (see Fig 11).

## Discussion

This study presents a model showing the effects of population age, time of day, and gathering location on the spread of the epidemic. Based on the proposed model Eqs (9-10), the non-trivial equilibria of the system is presented in Table 3. Based on Lemma 1, the equilibria do not take the form of the asymptotic case (Eq (11)) of the model and therefore are not stable. Although the proposed model is a simplification of the total complexity of the COVID-19 epidemic, the asymptotic solution without any intervention will result in a high percentage 39% children, 89% adults and 38.5% children, 100% adults of infected individuals as shown in Figs 3 and 6, respectively.

In addition, the asymptotic solutions of the model are insignificant for themselves as in practice these results occur after *A* days (13 years). The main epidemic dynamics as obtained from both the SIRD model Eqs (9-10) and the *hybrid* model is a few month long, as shown in Figs 3 and 6. Besides, the asymptotic solution can be used to easily obtain *D*_*a*_ and *D*_*c*_ as they accumulative sums of infected infected adults and children multiplied by a factor *ψ*_*a*_ and *ψ*_*c*_ and do not change after the epidemic is over. As a result, *α* = 2.1 *·* 10^*−*4^ is not used as for 82 days (as shown in Fig 3) only 0.017% of the children will become adults which effectually has only a minor affect on the epidemic results.

We presented the proposed model as a stochastic process using the Markov chain modeling approach as shown in Lemma 1. Using this representation, it is easy to find the stationary state of the chain (Eq (11)) and therefore the possible asymptotic forms of the SIRD model Eqs (9-10).

Furthermore, two policies for controlling the epidemic and preventing outbreak have been investigated using the *hybrid* model. First, the duration of working and schooling day has an influence (up to 10%) on the maximum of infected individuals but can prevent outbreaks under the assumption that during this time adults do not contact children at all as shown in Fig 7. These results match the conclusions reported by Keskinocak et al. (2020) for Georgia state [35]. In addition, these results match the results reported by Di Domenico [16] that school closure has a minor influence on the epidemic peak, just delays it. Second, a partial lockdown of adults and children show that adults and children lockdown has a similar first-order effect (linear coefficients of *L*_*a*_ and *L*_*c*_) but the combination between the two (*L*_*a*_*L*_*c*_) has a bigger weight on the average infection rate as shown in Eqs (19) and (21). These results match the conclusions reported by Aglar et al. (2020) for the state of Georgia regarding the voluntary lockdown with school closure [36]. By the same token, a lockdown of approximately half of the adult population prevents an outbreak where children lockdown has a less significant influence as shown in Figs 8 and 9. These results are based on values from Table 1, which can vary significantly between countries and depend on other hyperparameters such as population density, age distribution of the population and others.

The model explains the dynamics that took place between August 15 and September 15 (2020) in Israel as presented in Fig 11 with mean square error (MSE) of 0.205. The opening of the schools is equal to reducing the lockdown for children with some relaxation (smaller *L*_*a*_) of the lockdown for adults (because if the children in school they can go to work) with less then half (50%) of the adult population voluntary in lockdown. Also, the schooling hours were reduced to less than 6 hours each day. From Figs 7a and 8c it is easy to see that this policy predicts high increase in the infection rate, as indeed happened. Keeping the schools open while keeping the increase in the infection rate from increasing significantly is possible if the schooling hours are longer (8-9 hours) as shown in Fig 7a. The influence of this policy in Israel during the school opening which take place in September 1 show that the *R*_0_ can be reduced by 0.83 in comparison to a policy in which children go to school every other day for five hours, as shown in Fig 11. Also, if at least half of the adult population will be in lockdown the influence of the schools on the infection rate will be relatively small as shown in Fig 8b.

In the case of a future pandemic virus, researchers will be able to use our approach to predict the exact consequences of choosing “Lockdown” strategies, especially those needed for pandemics with a lack of immunity in the world’s population. Understanding the age-based dynamics in COVID-19 spread in the population will be crucial for the development of an optimal NPI policy, as well as for optimizing current polices and analyzing clinical data. The further extensions of the model will be used to learn how to manage international travel between countries with a range of healthcare system abilities in both the context of the COVID-19 epidemic and for other epidemics scenarios.

## Data Availability

All data available in the sources cited in the paper.

https://teddylazebnik.info/coronavirus-sir-simulation/index.html

## Notes

### Competing Interest Statement

The authors have declared no competing interest.

### Funding Statement

no external funding was received

